# Dietary exposures and common mental illness in the Netherlands Study of Depression and Anxiety (NESDA): a cohort-level GLAD project analysis

**DOI:** 10.64898/2026.02.05.26345645

**Authors:** Mariska Bot, Brenda W.J.H. Penninx

## Abstract

**Background:** Worldwide, common mental disorders such as anxiety and depression are major contributors to disability. However, the role of diet as a risk factor for anxiety and depression remains underexplored. Therefore, we investigated the associations between food groups and major depressive disorder (MDD) and anxiety disorders, following a harmonized protocol to enable integration of studies.

**Methods:** We analysed data from 1,634 participants in the Netherlands Study of Depression and Anxiety to examine cross-sectional associations between 14 dietary exposures—derived from a 238-item Food Frequency Questionnaire (fruit, vegetables, legumes, whole grains, nuts and seeds, milk, red meat, processed meat, sweet drinks, fibre, calcium, omega-3 fatty acids, polyunsaturated fatty acids, and trans fats)—and anxiety and depressive disorders in the past month (assessed with the Composite International Diagnostic Interview). Secondary outcomes were depressive symptoms (Quick Inventory of Depressive Symptomatology score ≥13 vs. <13) and anxiety symptoms (Beck Anxiety Index score ≥16 vs. <16). Logistic regression analyses were conducted for each dietary exposure, with depression and anxiety measures as outcomes.

**Results:** 8.7% had MDD and 14.4% had an anxiety disorder in the past month. Higher vegetable intake was associated with lower odds of depression and anxiety disorders. Additionally, higher intakes of omega-3 fatty acids, red meat, whole grains, and fibre were associated with lower odds of depression and anxiety, whereas higher intake of trans fats was associated with increased odds of these disorders. Other dietary exposures were not significantly related to depression or anxiety.

**Discussion:** Certain dietary exposures, particularly vegetables, as well as omega-3 fatty acids, red meat, whole grains, and fibre, were associated with depression and anxiety outcomes. These findings may contribute to integration of results in Global Burden of Diseases initiatives on exploring dietary risk factors of depression and anxiety.

## Introduction

The Global Burden of Diseases, Injuries, and Risk Factors Study (GBD) collects data on health trends worldwide and serve as an important tool for governments and policy makers to guide local, regional, and global health decisions. It is one of the world’s largest scientific collaborations and creates a unique platform to compare the prevalence of diseases, injuries and risk factors across age groups, sexes, countries, regions and times. The GBD 2021 study shows that common mental disorders like depressive and anxiety disorders are among the leading causes of disease (Diseases & Injuries, 2024); with depression and anxiety ranking as the second and sixth highest cause of years lived with disability (YLD), respectively. Furthermore, the disability adjusted life years (DALYs) also increased between 2010 and 2021 for both depression and anxiety. Because of the high burden related to depression and anxiety, it is critical to identify risk factors for these common mental disorder, which can assist in early prediction, prevention, and better treatment outcomes. One potential relevant set of risk factors that affects depression and anxiety are lifestyle factors. Lifestyle factors are modifiable risk factors, and may therefore provide an important target to predict, prevent and treat depression and anxiety. A meta-review on meta-analysis of prospective cohort studies, Mendelian randomization studies, and randomized controlled trials confirms that various lifestyle behaviors (e.g. diet quality, physical activity, smoking, and sleep) affects the risk of depression and anxiety (Firth et al., 2020).

Furthermore, in a France study, it was estimated that 14% of incident cases of depression in adults were attributable to a combination of poor lifestyle factors (unhealthy diet, weight, and smoking) (Adjibade et al., 2018). Despite evidence showing the contributions of lifestyle exposures to these common mental disorders, the GBD does not currently generate these lifestyle exposure-mental disorder outcome pairings. This is largely due to difficulties determining directionality and causality, and difficulties operationalizing lifestyle factors in a consistent way across studies. In addition, current studies on the role of lifestyle risk factors for common mental disorders (CMD) like depression and anxiety often have small sample sizes, inconsistent methods, and lack of global representation (Ashtree et al., 2025). To address these limitations, the Global burden of disease Lifestyle And mental Disorder (GLAD) Taskforce was established (Ashtree et al., 2025). The GLAD Taskforce is a large-scale, coordinated, international collaborative effort that collaborates with the GBD on the comprehensive evaluation of the association of lifestyle risk factors with CMDs. The first GLAD project will work specifically on the association of diet with anxiety and depression. GLAD developed a harmonized data analysis protocol that can be followed by individual studies. After combining results of individual studies, the project aims to generate robust, comparable evidence on the global risk of CMDs attributable to lifestyle factors, ultimately enabling their inclusion as risk factors for CMDs within the GBD framework.

Here, we analysed data of a large naturalistic cohort study on depressive and anxiety disorders (Netherlands Study on Depression and Anxiety; NESDA), and aimed to study the association of dietary factors with depressive and anxiety disorders. This extends previous work of Gibson-Smith et al. (Gibson-Smith et al., 2020), who used NESDA data to study associations between food groups and depression and anxiety outcomes. Because the analyses in Gibson-Smith were different from those proposed by the GLAD protocol, these analyses were redone in line with the GLAD protocol. We hypothesize that more healthful lifestyle factors, such as higher intakes of healthful dietary components (e.g. fruits, vegetables, whole grains, and fiber) will be associated with a lower risk of CMDs. Conversely, we hypothesize that less healthful lifestyle factors, such as higher intake of less healthful dietary components (e.g. processed meat, sugar-sweetened beverages) will be associated with a higher risk of CMDs.

## Methods

### Design

Data from the Netherlands Study of Depression and Anxiety (NESDA) were used (Penninx et al., 2021). NESDA is an ongoing longitudinal cohort study which aimed to investigate the course and consequences of having depressive and anxiety disorders. The baseline sample consists of 2981 participants (aged 18-65 years) with and without depressive and anxiety disorders.

Baseline and follow-up (after 2, 4, 6, 9, 13 years) in-depth interviews were done in which mental health status, anthropometric measurements, biological measurements and lifestyle factors were assessed. A new round of follow-up assessments is currently ongoing. At the 9-year assessment, a food frequency questionnaire (FFQ) was included. All participants completed written informed consent forms and the research protocol was approved by the Ethical Committee of the participating universities.

We selected all participants that completed the 9-year FFQ and excluded those with extreme energy intake (men: <800 or >4000 Kcal/day, women: <500 or 3500 Kcal/day). This resulted in 1634 participants; see Figure 1.

**Figure 1.**
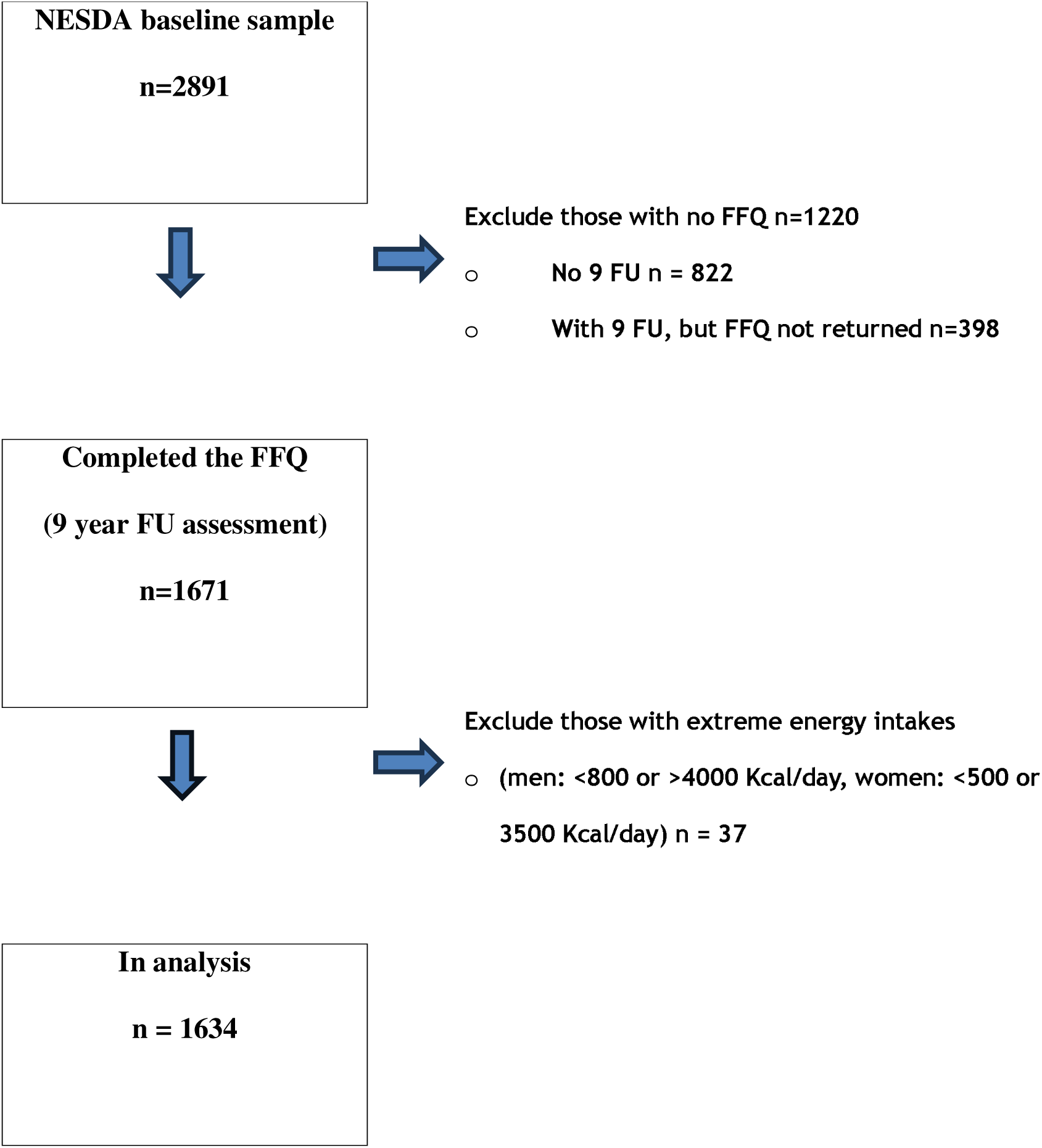
Flow chart NESDA. Abbreviations: FFQ: Food Frequency Questionnaire, FU: Follow-up.

### FFQ

At the 9-year follow-up assessment, dietary intake was assessed with a 238-item, semi-quantitative FFQ (Gibson-Smith et al., 2020). The FFQ asked about the frequency, amount, and type of food consumed in the past month. Using the 2014 Dutch Food Composition Table, the daily intakes (g/day) of the 238 food items were calculated. For missing amounts of reported food items, population medians were used. The FFQ also included the option to add food items consumed within the past week that were not included in the questionnaire. These items were manually re-categorised to comparable food items where possible. Each manual adjustment was made after consensus by two nutritional scientists.

The GBD includes 15 dietary exposures that are related to physical health outcomes, and the GLAD consortium lists an additional exposure (ultra-processed food) due to its relationship with mental disorders (Lane et al., 2024), bringing the total to sixteen dietary exposures. Fourteen of these could be derived from the FFQ in NESDA: Fruit (g/day), Vegetables (g/day), Legumes (g/day), Whole grains (g/day), Nuts and seeds (g/day), Milk (g/day), Red meat (g/day), Processed meat (g/day), Sweet drinks (g/day), Fibre (g/day), Calcium (g/day), Omega-3 fatty acids (mg/day), Poly Unsaturated Fatty Acids (PUFA) (% of total energy), and Transfat (% of total energy). In contrast, Sodium and Ultra-processed food could not be calculated in NESDA.

### Depression and anxiety

At each interview, the presence of a Diagnostic and Statistical Manual of Mental Disorders, Fourth Edition (DSM-IV) major depressive disorder (MDD) or anxiety disorder (social phobia, agoraphobia, general anxiety disorder and panic disorder) was assessed using the Composite International Diagnostic Interview (CIDI) version 2.1 (Wittchen, 1994).

Longitudinal incident analysis were not feasible, given the small number of new onset of MDD or anxiety cases between year 9-13 (i.e. after the FFQ assessment in year 9) in participants without lifetime CMD in year 9 (n=8 and n=8, respectively). Therefore, we measured the presence of MDD and anxiety disorders (past month diagnosis) in the 9-year follow-up.

At each follow-up, the Inventory of Depressive Symptomatology self-report version (IDS-SR) (Rush et al., 1996) and Beck Anxiety Inventory (BAI) (Beck et al., 1988) were used to measure the severity of depression and anxiety, respectively. Using the 9-year follow-up IDS-SR, we generated the Quick IDS scores (QIDS)(Rush et al., 2003), and classified participants with (>=13) and without (<13) depressive symptoms in line with the GLAD protocol (Ashtree et al., 2025). Similarly, with the 9-year follow-up BAI we classified participants with (>=16) and without (<16) anxiety symptoms (Ashtree et al., 2025).

### Covariates

Sex, age, years of education were measured at 9 years follow-up. In addition, we calculated the presence of lifetime MDD or anxiety disorders, using the CIDI interviews of previous measurements.

Furthermore, the following 9-year follow-up lifestyle variables were included: current smoking (yes/no), measured body mass index (BMI; kg/m^2^) and physical activity. Physical activity during the past week was measured with the International Physical Activity Questionnaire (IPAQ) and expressed as 1000 MET min/week (Craig et al., 2003).

### Statistical analysis

Analysis were done with SPSS and R version (4.4.3). First, descriptive data were given for the variables included in the analyses (Table 1). Next, we ran a series of logistic regression analyses linking the food groups (independent variables) to the depression and anxiety outcomes (dependent variables).

**Table 1.** NESDA sample characteristics n=1634.

|  |  |
| --- | --- |
| Sex n (%) |  |
| Male | 526 (32.2%) |
| Female | 1108 (67.8%) |
| Age in years, mean (SD) | 52.0 (13.2) |
| Education in years, mean (SD) | 13.2 (3.3) |
| Current smoker, n (%) | 380 (23.3%) |
| Physical Activity (total MET minutes/week), (median IQR)* | 2880 (3880) |
| Body Mass Index, kg/m<sup>2</sup>, mean (SD)* | 26.3 (4.8) |
| MDD in past month (9 year follow-up) n (%) | 142 (8.7%) |
| Anxiety disorder in past month (9 year follow-up) n (%) | 236 (14.4%) |
| Q-IDS $\geq 13$, n (%)* | 132 (8.2%) |
| BAI $\geq 16$, n (%)* | 235 (14.6%) |
| Fruit g/day, mean (SD) | 182.5 (137.1) |
| Vegetables g/day, mean (SD) | 178.5 (101.4) |
| Legumes g/day, median (IQR) | 12.5 (27.5) |
| Whole grains g/day, mean (SD) | 137.2 (75.2) |
| Nuts and seeds g/day, median (IQR) | 10.3 (20.3) |
| Milk g/day, median (IQR) | 114.3 (217.1) |
| Red meat g/day, mean (SD) | 43.6 (33.3) |
| Processed meat g/day, median (IQR) | 10.3 (19.2) |
| Sweet drinks g/day, median (IQR) | 15.6 (103.6) |
| Fibre g/day, mean (SD) | 23.7 (7.7) |
| Calcium g/day, mean (SD) | 1.1 (0.4) |
| Omega-3 (DHA and EPA) mg/day, median (IQR) | 179.6 (213.9) |
| PUFA (% energy), mean (SD) | 6.9 (2.2) |
| Transfat (% energy), mean (SD) | 0.4 (0.1) |
| Sodium g/day | NA** |
| Ultra-processed food g/day | NA** |

\*missing values: physical activity, n=117; body mass index, n=212; Q-IDS n=19, BAI n=22
\*\*NA=Not available in NESDA.
Abbreviations: BAI: Beck Anxiety Inventory, g: gram, IQR: interquartile range, MDD: major depressive disorder, PUFA: Polyunsaturated Fatty Acids, Q-IDS: Quick Inventory of Depressive Symptomatology, SD: standard deviation.

Separate logistic regression models were made for each of the 14 food groups with each of the CMD outcomes. The food groups were modelled as continuous variables (see Supplemental Table 5 for abbreviations). The dependent variables were A) MDD in the past month, B) anxiety disorder in the past month, C) QIDS >=13 (vs. <13) and D) BAI (>=16 vs. <16). MDD in the past month and anxiety disorder in the past month were the primary outcomes.

We created several models. The first model was an univariable model. The second model was adjusted for sex, age and education level. The third model was additionally adjusted for smoking, BMI and physical activity (with a smaller n of 1338 due to missing values). We also planned to run a fourth model additionally adjusted for the presence of lifetime MDD and anxiety disorders (yes/no), however, this resulted in extreme odds ratio’s for the lifetime CMD variables predicting current CMD (due to high overlap between lifetime disorders and current disorders). Therefore, the fourth model was omitted.

As sensitivity analyses, we accounted for differences in total energy intakes by adjusting the 14 food groups for energy intake using the Willett energy adjustment method (Willett et al., 1997). We regressed the food groups as dependent variables against total energy intake (kcal/day) as the independent variable, predicted the residuals from these models, and repeated the four models described above with the energy-adjusted residuals.

No adjustment for multiple testing was done, therefore, all analysis should be considered explorative.

For the primary outcome models, assumptions were checked. No outliers were observed and there was no indication of multicollinearity (all variance inflation factors were <10). The linearity of the logit assumption was checked by checking the significance of the interaction between the food group*ln(food group) in the logistic regression model. Violations of the linearity of the logit were observed for Fruit, Nuts and Seeds, and PUFA (for MDD), and Milk and Calcium (for anxiety disorders). Natural logarithmic (LN) transformations of the food groups Fruit, Nuts and Seeds, Milk, and Calcium yielded similar associations with CMD as the untransformed data; however, the normality of their distributions worsened after transformation. Therefore, the original (untransformed) data were used for these groups to enhance interpretability. In contrast, for PUFA, both univariable and fully adjusted analyses became significant for MDD after LN transformation, and the normality of the distribution improved. Consequently, the results of the LN-transformed analysis for PUFA are presented in Supplemental Table 6 as a sensitivity analysis.

## Results

Our sample predominantly consisted of females (67.8%), with an average age of 52 years. At the 9-year follow-up assessment, 8.7% had MDD and 14.4% had an anxiety disorder in the past month (Table 1).

### MDD past month

After adjusting for sex, age and education, every gram increase in vegetable intake was associated with a 0.998-fold lower odds of MDD (95%CI: 0.996-1.000) and every mg increase in omega-3 was associated with a 0.999-fold lower odds of MDD (95%CI: 0.997-1.000; Table 2). Other dietary exposures were not significantly related to MDD in these models. After additional adjustment for other lifestyle factors, none of the dietary exposure were related to MDD in the past month (Table 2).

**Table 2.**
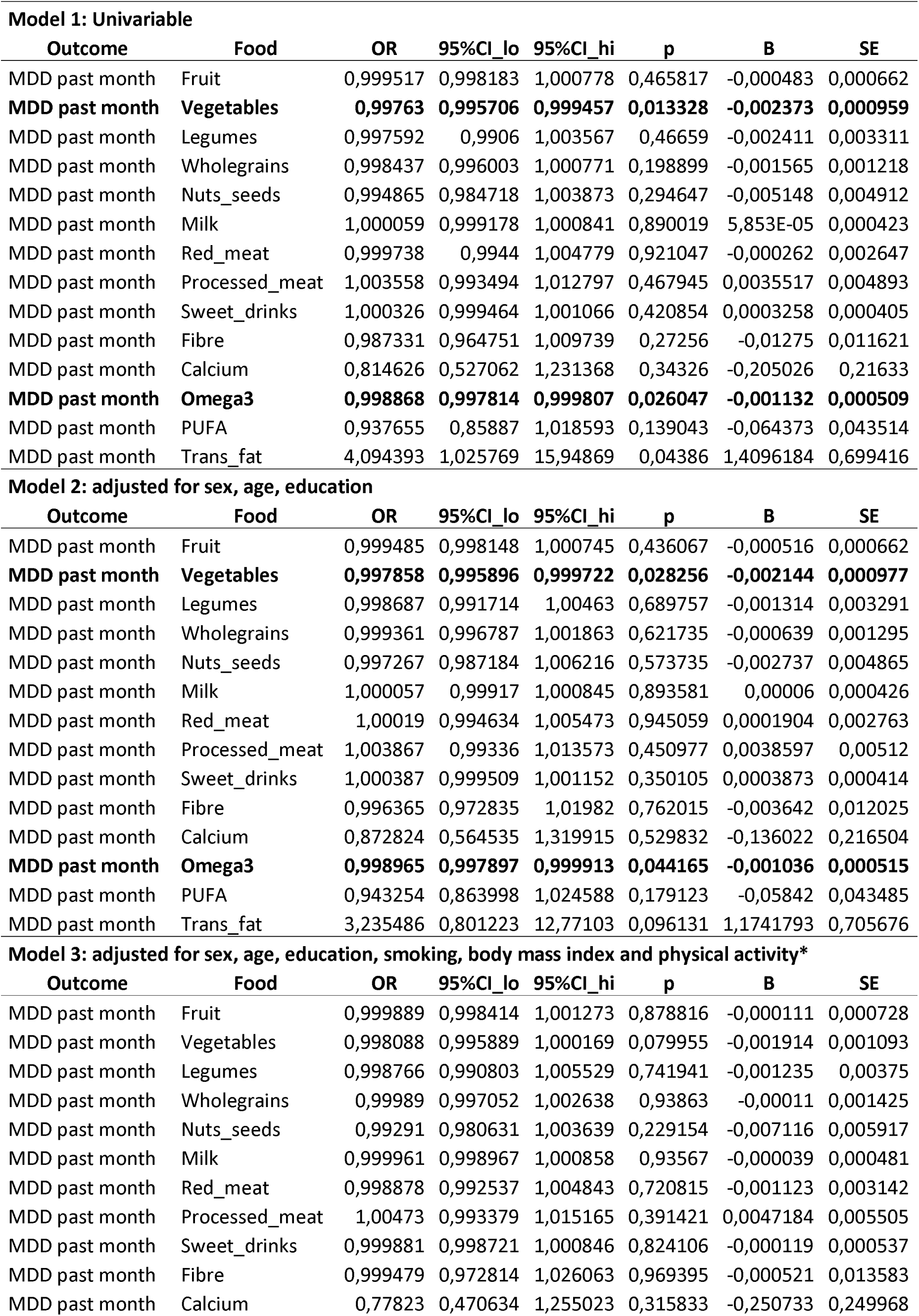
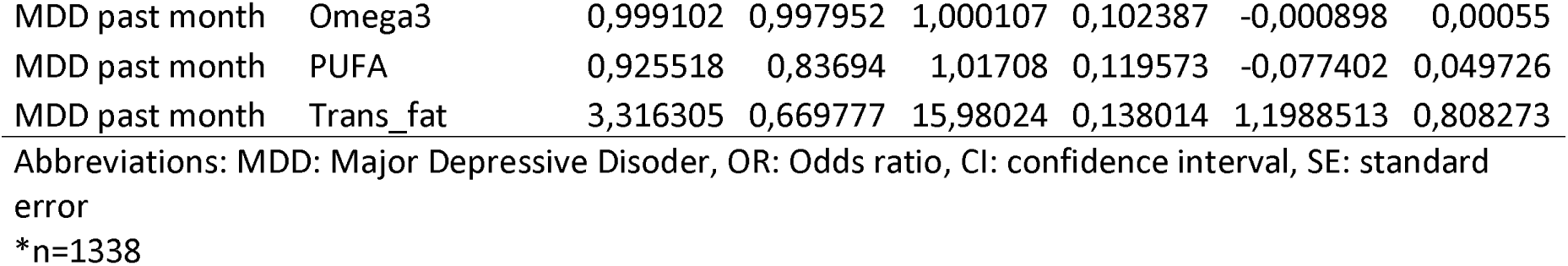
Logistic regression on the association of food groups with MDD in the past month (n=1634) Model 1: Univariable.

Sensitivity analyses accounting for total energy intake (Willet-adjusted model) generated largely similar patterns for the food groups: higher vegetable intake and higher omega-3 intake were related to a lower odds of MDD in the past month. In addition, transfat intake was significantly related to MDD in the model adjusted for sex, age and education (Supplemental table 1).

### Anxiety disorder past month

Higher vegetable intake was also consistently related with a lower odds of anxiety disorder in the past month (Table 3). Furthermore, higher omega-3 intake was related to a lower odds of anxiety disorder (Table 3). In the fully adjusted models, higher omega-3 intakes were no longer significantly related to anxiety disorders, but higher red meat intake became significantly related to a lower odds of anxiety disorder (Table 3).

**Table 3.** Logistic regression on the association of food groups with anxiety disorder in the past month (n=1634)

| **Model 1: Univariable** |  |  |  |  |  |  |  |
| --- | --- | --- | --- | --- | --- | --- | --- |
| **Outcome** | **Food** | **OR** | **95%CI_lo** | **95%CI_hi** | **p** | **B** | **SE** |
| Anx past month | Fruit | 0,9993348 | 0,9982656 | 1,0003603 | 0,2130327 | -0,0006654 | 0,0005343 |
| **Anx past month** | **Vegetables** | **0,9976244** | **0,9961033** | **0,999086** | **0,0018077** | **-0,0023785** | **0,0007623** |
| Anx past month | Legumes | 1,0001943 | 0,9952388 | 1,0046636 | 0,9353077 | 0,0001943 | 0,0023933 |
| Anx past month | Wholegrains | 0,9998071 | 0,9979407 | 1,0016222 | 0,8371458 | -0,0001929 | 0,0009387 |
| Anx past month | Nuts_seeds | 1,0005596 | 0,9934288 | 1,0071538 | 0,8726154 | 0,0005595 | 0,0034894 |
| Anx past month | Milk | 1,0003561 | 0,9996997 | 1,00097 | 0,2694752 | 0,000356 | 0,0003224 |
| Anx past month | Red_meat | 0,9966261 | 0,9921819 | 1,0008883 | 0,1292814 | -0,0033796 | 0,0022279 |
| Anx past month | Processed_meat | 1,0031495 | 0,9950953 | 1,0107709 | 0,4290456 | 0,0031446 | 0,0039763 |
| Anx past month | Sweet_drinks | 1,0005531 | 0,9999076 | 1,0011508 | 0,0787305 | 0,000553 | 0,0003145 |
| Anx past month | Fibre | 0,9961072 | 0,9782265 | 1,0139445 | 0,6696632 | -0,0039004 | 0,0091427 |
| Anx past month | Calcium | 1,0854527 | 0,7815037 | 1,4918231 | 0,6187132 | 0,0819972 | 0,1647605 |
| **Anx past month** | **Omega3** | **0,9989681** | **0,9981623** | **0,9997056** | **0,0087531** | **-0,0010325** | **0,0003938** |
| Anx past month | PUFA | 1,0008835 | 0,9378264 | 1,0654648 | 0,9783353 | 0,0008831 | 0,0325208 |
| Anx past month | Trans_fat | 1,1630284 | 0,3725316 | 3,5722994 | 0,7933 | 0,1510273 | 0,5763789 |
| **Model 2: adjusted for sex, age, education** |  |  |  |  |  |  |  |
| **Outcome** | **Food** | **OR** | **95%CI_lo** | **95%CI_hi** | **p** | **B** | **SE** |
| Anx past month | Fruit | 0,9994407 | 0,9983703 | 1,0004676 | 0,2954966 | -0,0005595 | 0,0005348 |
| **Anx past month** | **Vegetables** | **0,9979513** | **0,9963987** | **0,9994443** | **0,0083918** | **-0,0020508** | **0,000778** |
| Anx past month | Legumes | 1,0013804 | 0,9964749 | 1,0058308 | 0,5605829 | 0,0013794 | 0,0023702 |
| Anx past month | Wholegrains | 1,000472 | 0,9984873 | 1,0024234 | 0,6379293 | 0,0004719 | 0,0010027 |
| Anx past month | Nuts_seeds | 1,0026358 | 0,9954822 | 1,0092804 | 0,4519596 | 0,0026323 | 0,0034997 |
| Anx past month | Milk | 1,0003506 | 0,9996914 | 1,0009692 | 0,2793201 | 0,0003506 | 0,0003241 |
| Anx past month | Red_meat | 0,9962036 | 0,9915744 | 1,0006634 | 0,1020889 | -0,0038036 | 0,0023266 |
| Anx past month | Processed_meat | 1,003059 | 0,9946129 | 1,0110908 | 0,4648946 | 0,0030544 | 0,0041794 |
| Anx past month | Sweet_drinks | 1,0004329 | 0,9997586 | 1,0010618 | 0,1893736 | 0,0004328 | 0,0003297 |
| Anx past month | Fibre | 1,0032096 | 0,9845058 | 1,0219561 | 0,7363308 | 0,0032045 | 0,0095168 |
| Anx past month | Calcium | 1,1514135 | 0,8276037 | 1,5860346 | 0,3949309 | 0,1409904 | 0,1657327 |
| **Anx past month** | **Omega3** | **0,999166** | **0,9983601** | **0,9999026** | **0,033933** | **-0,0008343** | **0,0003934** |
| Anx past month | PUFA | 1,0059181 | 0,9420513 | 1,0713744 | 0,8571479 | 0,0059006 | 0,0327802 |
| Anx past month | Trans_fat | 0,9452155 | 0,2981785 | 2,9480349 | 0,9231538 | -0,0563423 | 0,5840891 |
| **Model 3: adjusted for sex, age, education, smoking, body mass index and physical activity\*** |  |  |  |  |  |  |  |
| **Outcome** | **Food** | **OR** | **95%CI_lo** | **95%CI_hi** | **p** | **B** | **SE** |
| Anx past month | Fruit | 0,9999345 | 0,9987468 | 1,0010717 | 0,9118851 | -6,552E-05 | 0,0005921 |
| **Anx past month** | **Vegetables** | **0,9981152** | **0,9963377** | **0,9998185** | **0,0338081** | **-0,0018866** | **0,0008889** |
| Anx past month | Legumes | 1,0009061 | 0,9950323 | 1,0061852 | 0,7485624 | 0,0009057 | 0,0028257 |
| Anx past month | Wholegrains | 1,0006955 | 0,9984613 | 1,0028867 | 0,5371785 | 0,0006953 | 0,0011267 |
| Anx past month | Nuts_seeds | 1,0020344 | 0,9938144 | 1,0095407 | 0,6102458 | 0,0020324 | 0,0039872 |
| Anx past month | Milk | 1,0002956 | 0,9995401 | 1,0010053 | 0,4275891 | 0,0002955 | 0,0003725 |
| **Anx past month** | **Red_meat** | **0,9935747** | **0,9881315** | **0,9987776** | **0,0183361** | **-0,006446** | **0,0027328** |
| Anx past month | Processed_meat | 1,0031899 | 0,9939036 | 1,0119778 | 0,4867165 | 0,0031848 | 0,0045789 |
| Anx past month | Sweet_drinks | 1,000221 | 0,999399 | 1,0009646 | 0,5767116 | 0,000221 | 0,000396 |
| Anx past month | Fibre | 1,0028613 | 0,9813565 | 1,0244148 | 0,7940147 | 0,0028572 | 0,0109429 |
| Anx past month | Calcium | 1,1781469 | 0,8036277 | 1,7085501 | 0,3935863 | 0,1639428 | 0,1921658 |
| Anx past month | Omega3 | 0,9992988 | 0,9984173 | 1,000093 | 0,1006115 | -0,0007014 | 0,0004272 |
| Anx past month | PUFA | 1,0272566 | 0,9543438 | 1,1025171 | 0,4643912 | 0,0268917 | 0,0367557 |
| Anx past month | Trans_fat | 0,8676411 | 0,2276166 | 3,2385378 | 0,8338183 | -0,1419771 | 0,6767032 |

Abbreviations: Anx: Anxiety Disorder, OR: Odds ratio, CI: confidence interval, SE: standard error
\*n=1338

Sensitivity analyses accounting for total energy intake (Willet-adjusted model) generated similar patterns for the food groups: vegetable intake and omega-3 intake were related to less anxiety in (most) models. Red meat intake was also related to less anxiety in all three models (Supplemental Table 2).

### QIDS depression

When using the QIDS as an indicator of depression, vegetable intake was significantly related to QIDS depression in the univariable model and after adjustment for sex, age and education (Table 4); and borderline significant in the fully adjusted model. We also observed additional significant associations for QIDS depression with wholegrains, fibre and trans fat in the univariable model and after adjustment for sex, age and education (Table 4). After additional adjustment for lifestyle factors, only transfat remained associated with QIDS depression (Table 4), with higher transfat intake related to increased QIDS depression.

**Table 4.**
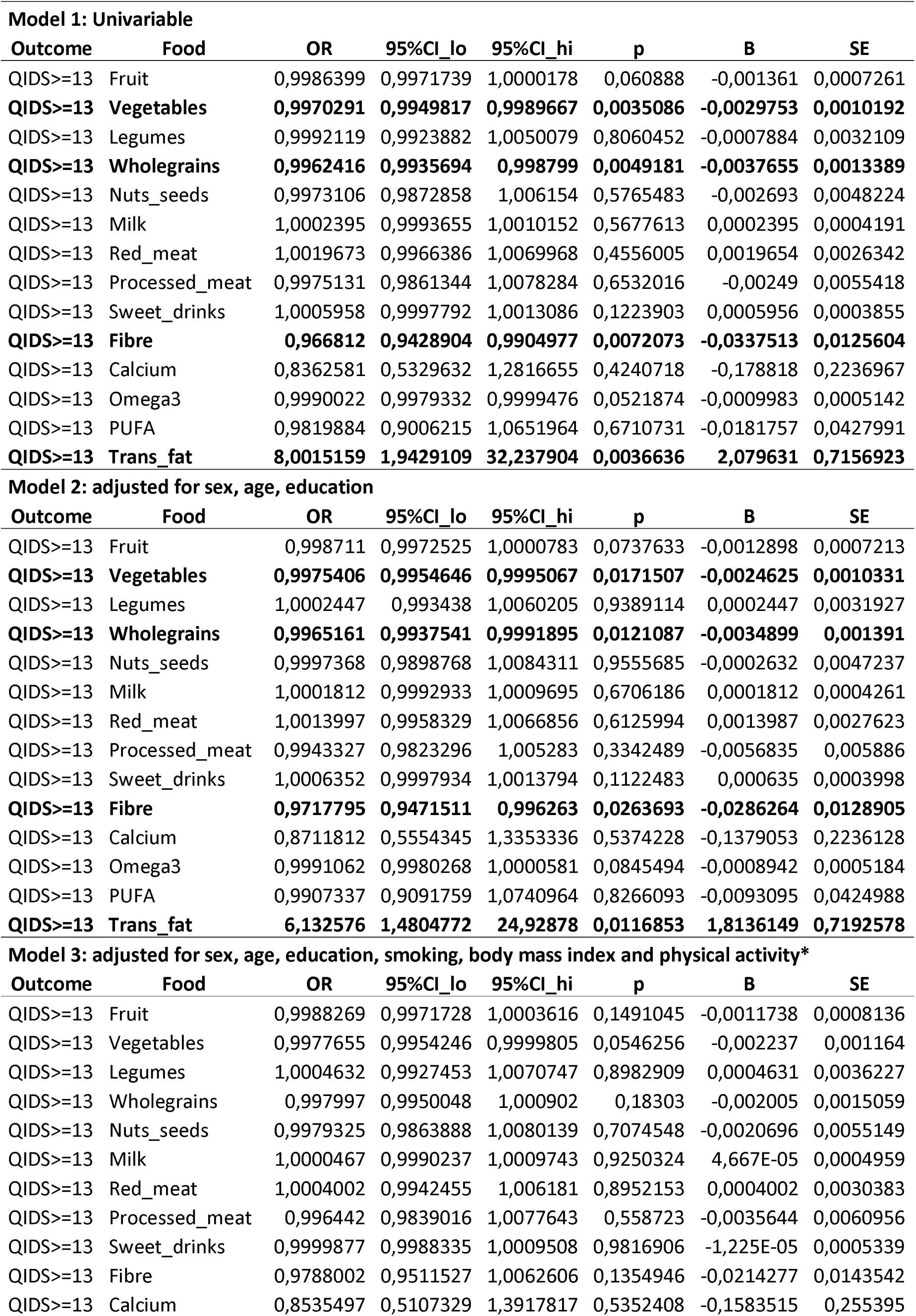
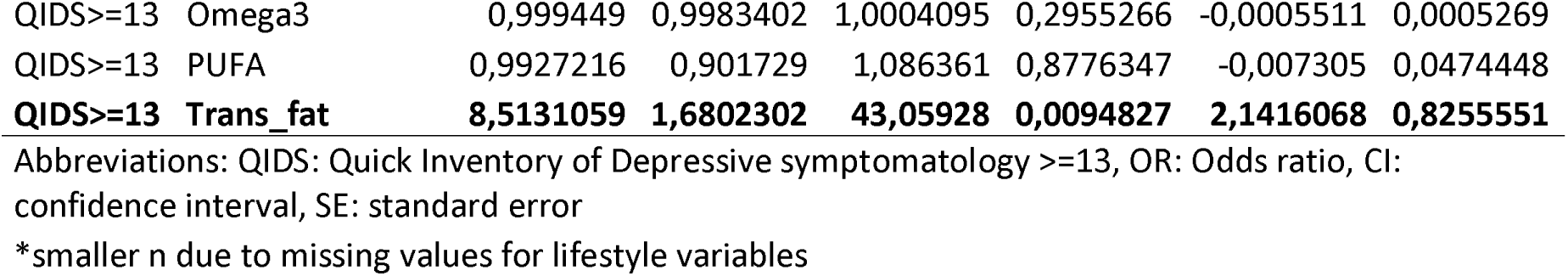
Logistic regression on the association of food groups with QIDS depression (n=1615) Model 1: Univariable.

Sensitivity analyses accounting for total energy intake (Willet-adjusted model) generated similar patterns for the food groups: vegetable intake was related to a lower odds of QIDS depression in all models (Supplemental table 3). We also observed additional significant associations for QIDS depression with wholegrains, fibre and trans fat in various models (Supplemental Table 3).

### BAI anxiety

The only significant food group related to BAI anxiety was vegetable intake in the univariable model and after adjustment for sex, age and education (Table 5), with higher intakes of vegetables being related to a lower odds of anxiety.

**Table 5.** Logistic regression on the association of food groups with BAI anxiety (n=1612) Model 1: Univariable.

| Outcome | Food | OR | 95%CI_lo | 95%CI_hi | p | B | SE |
| --- | --- | --- | --- | --- | --- | --- | --- |
| BAI>=16 | Fruit | 0,9997614 | 0,9987177 | 1,0007645 | 0,6474873 | -0,0002386 | 0,0005218 |
| **BAI>=16** | **Vegetables** | **0,9976544** | **0,9961284** | **0,9991204** | **0,0021325** | **-0,0023483** | **0,0007646** |
| BAI>=16 | Legumes | 0,9992162 | 0,9940203 | 1,0038756 | 0,7544834 | -0,0007841 | 0,0025074 |
| BAI>=16 | Wholegrains | 0,9986482 | 0,9967195 | 1,0005187 | 0,1631706 | -0,0013527 | 0,0009701 |
| BAI>=16 | Nuts_seeds | 0,997573 | 0,990015 | 1,0045125 | 0,5115205 | -0,0024299 | 0,0037015 |
| BAI>=16 | Milk | 1,0000871 | 0,9993967 | 1,0007255 | 0,7965076 | 8,714E-05 | 0,0003379 |
| BAI>=16 | Red_meat | 1,0022186 | 0,9981063 | 1,0061961 | 0,281026 | 0,0022162 | 0,0020558 |
| BAI>=16 | Processed_meat | 1,0047965 | 0,996888 | 1,0123124 | 0,2204052 | 0,004785 | 0,0039047 |
| BAI>=16 | Sweet_drinks | 1,0004046 | 0,9997344 | 1,0010168 | 0,212614 | 0,0004045 | 0,0003245 |
| BAI>=16 | Fibre | 0,9934684 | 0,9754561 | 1,0114259 | 0,4778115 | -0,006553 | 0,0092318 |
| BAI>=16 | Calcium | 0,8990583 | 0,6390635 | 1,2496642 | 0,5336251 | -0,1064074 | 0,1709406 |
| BAI>=16 | Omega3 | 0,9994144 | 0,9986697 | 1,000097 | 0,1074195 | -0,0005858 | 0,0003639 |
| BAI>=16 | PUFA | 0,9998219 | 0,9366829 | 1,0644664 | 0,9956386 | -0,0001782 | 0,0325923 |
| BAI>=16 | Trans_fat | 1,8429657 | 0,5911475 | 5,6632902 | 0,2885454 | 0,6113761 | 0,5760537 |

**Model 2: adjusted for sex, age, education**
| Outcome | Food | OR | 95%CI_lo | 95%CI_hi | p | B | SE |
| --- | --- | --- | --- | --- | --- | --- | --- |
| BAI>=16 | Fruit | 0,9998358 | 0,9987948 | 1,0008376 | 0,7524135 | -0,0001642 | 0,0005206 |
| **BAI>=16** | **Vegetables** | **0,998165** | **0,996614** | **0,9996571** | **0,0181135** | **-0,0018367** | **0,0007772** |
| BAI>=16 | Legumes | 1,0006865 | 0,9955157 | 1,0053452 | 0,7831414 | 0,0006862 | 0,0024933 |
| BAI>=16 | Wholegrains | 0,9993945 | 0,9973534 | 1,0013942 | 0,5567749 | -0,0006057 | 0,0010307 |
| BAI>=16 | Nuts_seeds | 1,0003339 | 0,9928365 | 1,0072486 | 0,9274427 | 0,0003339 | 0,0036665 |
| BAI>=16 | Milk | 1,0000491 | 0,9993498 | 1,0006957 | 0,8859758 | 4,908E-05 | 0,0003422 |
| BAI>=16 | Red_meat | 1,0021306 | 0,9978138 | 1,0063201 | 0,3250153 | 0,0021284 | 0,0021625 |
| BAI>=16 | Processed_meat | 1,0036029 | 0,9952676 | 1,0115549 | 0,3836069 | 0,0035965 | 0,0041278 |
| BAI>=16 | Sweet_drinks | 1,0003604 | 0,9996664 | 1,0010009 | 0,2859247 | 0,0003604 | 0,0003377 |
| BAI>=16 | Fibre | 1,0025479 | 0,9836689 | 1,0214736 | 0,7912343 | 0,0025446 | 0,0096129 |
| BAI>=16 | Calcium | 0,9556515 | 0,6784069 | 1,3307201 | 0,791641 | -0,045362 | 0,1717085 |
| BAI>=16 | Omega3 | 0,9995882 | 0,9988406 | 1,0002736 | 0,2591072 | -0,0004119 | 0,000365 |
| BAI>=16 | PUFA | 1,008262 | 0,944407 | 1,0737135 | 0,8013199 | 0,008228 | 0,0326978 |
| BAI>=16 | Trans_fat | 1,3649909 | 0,4312497 | 4,2584512 | 0,5939711 | 0,3111478 | 0,5836681 |

**Model 3: adjusted for sex, age, education, smoking, body mass index and physical activity\***
| Outcome | Food | OR | 95%CI_lo | 95%CI_hi | p | B | SE |
| --- | --- | --- | --- | --- | --- | --- | --- |
| BAI>=16 | Fruit | 1,0002586 | 0,999093 | 1,0013808 | 0,6569746 | 0,0002585 | 0,0005821 |
| BAI>=16 | Vegetables | 0,9988497 | 0,9970981 | 1,0005357 | 0,1894227 | -0,001151 | 0,0008771 |
| BAI>=16 | Legumes | 1,0004175 | 0,994303 | 1,0059257 | 0,8873247 | 0,0004174 | 0,0029456 |
| BAI>=16 | Wholegrains | 1,0013816 | 0,99908 | 1,0036561 | 0,2357566 | 0,0013806 | 0,0011644 |
| BAI>=16 | Nuts_seeds | 0,998999 | 0,9901835 | 1,0070383 | 0,8155589 | -0,0010015 | 0,0042933 |
| BAI>=16 | Milk | 0,9999118 | 0,9990848 | 1,0006822 | 0,828126 | -8,825E-05 | 0,0004065 |
| BAI>=16 | Red_meat | 1,0014152 | 0,9965114 | 1,0061396 | 0,5633722 | 0,0014142 | 0,0024474 |
| BAI>=16 | Processed_meat | 1,0049892 | 0,9959137 | 1,0136784 | 0,2674538 | 0,0049768 | 0,0044879 |
| BAI>=16 | Sweet_drinks | 1,000056 | 0,9991736 | 1,0008415 | 0,8942079 | 5,603E-05 | 0,0004214 |
| BAI>=16 | Fibre | 1,0122181 | 0,9904754 | 1,0341374 | 0,2691891 | 0,0121441 | 0,0109908 |
| BAI>=16 | Calcium | 0,8932952 | 0,5959482 | 1,3209063 | 0,5779391 | -0,1128382 | 0,2028014 |
| BAI>=16 | Omega3 | 0,9998693 | 0,9990843 | 1,0005842 | 0,7318297 | -0,0001307 | 0,0003813 |
| BAI>=16 | PUFA | 1,0170483 | 0,9436452 | 1,0929327 | 0,6512697 | 0,0169047 | 0,0373998 |
| BAI>=16 | Trans_fat | 1,3657787 | 0,3591699 | 5,1174871 | 0,6451296 | 0,3117247 | 0,6768699 |

Abbreviations: BAI: Becks Anxiety Index >=16, OR: Odds ratio, CI: confidence interval, SE: standard error
\*smaller n due to missing values for lifestyle variables

Sensitivity analyses accounting for total energy intake (Willet-adjusted model) generated similar patterns for the food groups, but identified some additional food groups being related to BAI across various models (whole grains, fibre, calcium) (Supplemental Table 4).

## Discussion

Using the GLAD protocol on the comprehensive evaluation of the association of lifestyle risk factors with CMDs and building on previous work of Gibson-Smith et al., (2020) we studied associations between individual food groups and depression and anxiety disorders. Most consistent findings were observed for higher vegetable intake and reduced odds for depression and anxiety. In addition, omega-3 fatty acids, red meat, whole grain and fiber intakes were inversely related to CMDs in multiple models, and higher transfat intake was associated with increased odds of CMDs in multiple models.

Studies investigating the relationship of individual food groups with CMD generated mixed results. This is partly the result of variation in the definition of food groups across studies, which makes direct comparisons difficult. Generally, the direction of the relationships of the food groups was in line with our expectations. One exception was red meat intake, for we observed a protective association with CMD. This was also observed in Gibson-Smith et al. (2020) using the same NESDA dataset, and also in a different study, but in women only (Mikolajczyk et al., 2009). Multiple previous studies report on the association of vegetables with (incident) depression (Matison et al., 2021). Furthermore, similar to our study, a systematic review showed that wholegrain intake has been related to less depression and anxiety (Ross et al., 2023). Also, in line with our observations, omega-3 intake has been linked to reduced risk of maintenance of depressive episodes(Chaves et al., 2022).

Strengths of this study include its focus on both depression and anxiety disorders—two prevalent and highly comorbid mental health conditions—the use of a standardized protocol for calculating food groups and conducting statistical analyses, and the inclusion of a sample in which depression and anxiety were oversampled, thereby increasing statistical power.

However, several limitations should be noted. First, although NESDA has a longitudinal design, our analyses were primarily cross-sectional, as the FFQ was only administered at the 9-year follow-up. This observational design limits our ability to draw conclusions regarding temporal relationships. Second, dietary intake was assessed using a self-report FFQ, which may be subject to misreporting and recall bias; these biases may differ between individuals with and without common mental disorders. Third, given the multiple analyses specified in the GLAD protocol and lack of multiple testing correction, the results should be considered exploratory. Fourth, for certain food groups, the linearity of the logit assumption may have been slightly violated (for MDD: fruit, nuts/seeds, and PUFA; for anxiety: milk and calcium), and variable transformation did not resolve this issue. Therefore, findings related to these food groups should be interpreted with caution.

In summary, we found consistent associations between higher vegetable intake and reduced odds of depression and anxiety. Additionally, intakes of omega-3 fatty acids, red meat, whole grains, and fibre were inversely associated with common mental disorders (CMDs), whereas higher intake of trans fats was associated with increased odds of CMDs. By adhering to a standardized protocol, this study contributes to the generation of robust and comparable evidence regarding the global risk of CMDs attributable to lifestyle factors.

## Acknowledgement

The infrastructure for the NESDA study (www.nesda.nl) is funded through the Geestkracht program of the Netherlands Organisation for Health Research and Development (ZonMw, grant number 10-000- 1002) and financial contributions by participating universities and mental health care organizations (VU University Medical Center, GGZ inGeest, Leiden University Medical Center, Leiden University, GGZ Rivierduinen, University Medical Center Groningen, University of Groningen, Lentis, GGZ Friesland, GGZ Drenthe, Rob Giel Onderzoekscentrum).

## Data availability

All data produced in the present study are available upon reasonable request to the authors

