## Supplemental Tables for "Dietary exposures and common mental illness in the Netherlands Study of Depression and Anxiety (NESDA): a cohort-level GLAD project analysis"

| **Supplemental table 1. Logistic regression on the association of food groups with MDD in the past month, Willet adjusted model (n=1634)** | | | | | | | |
| --- | --- | --- | --- | --- | --- | --- | --- |
| **Model 1: Univariable** | | |  |  |  |  |  |
| **Outcome** | **Food** | **OR** | **95%CI_lo** | **95%CI_hi** | **p** | **B** | **SE** |
| MDD past month | Fruit_en | 0,9995 | 0,998162 | 1,000763 | 0,450535 | -0,000501 | 0,000663 |
| **MDD past month** | **Vegetables_en** | **0,99754** | **0,995593** | **0,999385** | **0,011049** | **-0,002463** | **0,000969** |
| MDD past month | Legumes_en | 0,99727 | 0,990141 | 1,003366 | 0,418739 | -0,002734 | 0,003381 |
| MDD past month | Wholegrains_en | 0,997553 | 0,994833 | 1,000223 | 0,075629 | -0,00245 | 0,001379 |
| MDD past month | Nuts_seeds_en | 0,993222 | 0,982501 | 1,002918 | 0,195344 | -0,006801 | 0,005252 |
| MDD past month | Milk_en | 1,000011 | 0,999104 | 1,000821 | 0,980089 | 1,092E-05 | 0,000438 |
| MDD past month | Red_meat_en | 0,999172 | 0,993504 | 1,004568 | 0,769333 | -0,000829 | 0,002826 |
| MDD past month | Processed_meat_en | 1,003098 | 0,992531 | 1,012867 | 0,549029 | 0,0030931 | 0,005162 |
| MDD past month | Sweet_drinks_en | 1,000297 | 0,999405 | 1,001061 | 0,478442 | 0,0002966 | 0,000419 |
| MDD past month | Fibre_en | 0,970248 | 0,940521 | 1,0001 | 0,053951 | -0,030204 | 0,015672 |
| MDD past month | Calcium_en | 0,674041 | 0,397868 | 1,116816 | 0,134137 | -0,394464 | 0,26333 |
| **MDD past month** | **Omega3_en** | **0,998809** | **0,997738** | **0,999762** | **0,021085** | **-0,001192** | **0,000517** |
| MDD past month | PUFA_en | 0,93543 | 0,856292 | 1,016675 | 0,127528 | -0,066749 | 0,043801 |
| **MDD past month** | **Trans_fat_en** | **4,014008** | **1,002758** | **15,66528** | **0,047292** | **1,3897903** | **0,700613** |
| **Model 2: adjusted for sex, age, education** | | | | |  |  |  |
| **Outcome** | **Food** | **OR** | **95%CI_lo** | **95%CI_hi** | **p** | **B** | **SE** |
| MDD past month | Fruit_en | 0,99944 | 0,998095 | 1,000708 | 0,400347 | -0,00056 | 0,000666 |
| **MDD past month** | **Vegetables_en** | **0,997648** | **0,995641** | **0,999551** | **0,018452** | **-0,002355** | **0,000999** |
| MDD past month | Legumes_en | 0,997985 | 0,990748 | 1,004143 | 0,555266 | -0,002017 | 0,003419 |
| MDD past month | Wholegrains_en | 0,998143 | 0,995342 | 1,000906 | 0,191132 | -0,001859 | 0,001422 |
| MDD past month | Nuts_seeds_en | 0,994279 | 0,983253 | 1,004205 | 0,286415 | -0,005737 | 0,005382 |
| MDD past month | Milk_en | 0,999934 | 0,999005 | 1,000761 | 0,882363 | -6,63E-05 | 0,000448 |
| MDD past month | Red_meat_en | 0,998822 | 0,992949 | 1,004438 | 0,688063 | -0,001179 | 0,002935 |
| MDD past month | Processed_meat_en | 1,00207 | 0,991017 | 1,012328 | 0,702536 | 0,0020681 | 0,005415 |
| MDD past month | Sweet_drinks_en | 1,000286 | 0,999369 | 1,001079 | 0,507917 | 0,0002859 | 0,000432 |
| MDD past month | Fibre_en | 0,974716 | 0,943882 | 1,005678 | 0,113488 | -0,025609 | 0,016181 |
| MDD past month | Calcium_en | 0,637054 | 0,368479 | 1,07325 | 0,098269 | -0,450901 | 0,272728 |
| **MDD past month** | **Omega3_en** | **0,998831** | **0,997728** | **0,999812** | **0,028021** | **-0,001169** | **0,000532** |
| MDD past month | PUFA_en | 0,938283 | 0,85853 | 1,020091 | 0,147521 | -0,063704 | 0,043984 |
| MDD past month | Trans_fat_en | 3,063147 | 0,754422 | 12,14097 | 0,113902 | 1,1194428 | 0,708106 |
| **Model 3: adjusted for sex, age, education, smoking, body mass index and physical activity*** | | | | | | | |
| **Outcome** | **Food** | **OR** | **95%CI_lo** | **95%CI_hi** | **p** | **B** | **SE** |
| MDD past month | Fruit_en | 0,999861 | 0,998378 | 1,001252 | 0,849641 | -0,000139 | 0,000732 |
| MDD past month | Vegetables_en | 0,997939 | 0,995698 | 1,000055 | 0,063788 | -0,002063 | 0,001113 |
| MDD past month | Legumes_en | 0,998253 | 0,990036 | 1,005227 | 0,65167 | -0,001749 | 0,003874 |
| MDD past month | Wholegrains_en | 0,999152 | 0,996083 | 1,002173 | 0,585432 | -0,000848 | 0,001555 |
| MDD past month | Nuts_seeds_en | 0,990124 | 0,977103 | 1,001786 | 0,119633 | -0,009925 | 0,006377 |
| MDD past month | Milk_en | 0,999872 | 0,99884 | 1,000807 | 0,798182 | -0,000128 | 0,000501 |
| MDD past month | Red_meat_en | 0,997825 | 0,991189 | 1,004118 | 0,510405 | -0,002178 | 0,003309 |
| MDD past month | Processed_meat_en | 1,003729 | 0,991914 | 1,014646 | 0,517871 | 0,0037222 | 0,005756 |
| MDD past month | Sweet_drinks_en | 0,999776 | 0,998573 | 1,000779 | 0,688759 | -0,000224 | 0,000559 |
| MDD past month | Fibre_en | 0,985932 | 0,951098 | 1,020916 | 0,433 | -0,014168 | 0,01807 |
| MDD past month | Calcium_en | 0,576476 | 0,309103 | 1,045588 | 0,076278 | -0,550821 | 0,310724 |
| MDD past month | Omega3_en | 0,999018 | 0,99784 | 1,000047 | 0,080956 | -0,000983 | 0,000563 |
| MDD past month | PUFA_en | 0,921684 | 0,832661 | 1,013737 | 0,104223 | -0,081552 | 0,050195 |
| MDD past month | Trans_fat_en | 3,187266 | 0,641203 | 15,39785 | 0,152353 | 1,1591636 | 0,809886 |
| Abbreviations: MDD: Major Depressive Disoder, OR: Odds ratio, CI: confidence interval, SE: standard error | | | | | | | |
| *n=1338 |  |  |  |  |  |  |  |

| **Supplemental tabel 2. Logistic regression on the association of food groups with anxiety disorder in the past month, Willet adjusted model (n=1634)** | | | | | | | |
| --- | --- | --- | --- | --- | --- | --- | --- |
| **Model 1: Univariable** | |  |  |  |  |  |  |
| **Outcome** | **Food** | **OR** | **95%CI_lo** | **95%CI_hi** | **p** | **B** | **SE** |
| Anx past month | Fruit_en | 0,9993012 | 0,9982288 | 1,0003296 | 0,1920981 | -0,000699 | 0,0005359 |
| **Anx past month** | **Vegetables_en** | **0,9974842** | **0,995943** | **0,9989634** | **0,0011032** | **-0,0025189** | **0,000772** |
| Anx past month | Legumes_en | 0,9997729 | 0,994694 | 1,0043406 | 0,9261806 | -0,0002272 | 0,0024518 |
| Anx past month | Wholegrains_en | 0,998969 | 0,9968287 | 1,0010768 | 0,3415467 | -0,0010315 | 0,0010846 |
| Anx past month | Nuts_seeds_en | 0,9989794 | 0,9912706 | 1,0061457 | 0,7877605 | -0,0010211 | 0,0037928 |
| Anx past month | Milk_en | 1,0002864 | 0,9996049 | 1,0009232 | 0,3923621 | 0,0002864 | 0,0003348 |
| **Anx past month** | **Red_meat_en** | **0,995119** | **0,9903915** | **0,9996867** | **0,0400632** | **-0,004893** | **0,0023832** |
| Anx past month | Processed_meat_en | 1,0019107 | 0,9933937 | 1,0099941 | 0,6509343 | 0,0019089 | 0,0042188 |
| Anx past month | Sweet_drinks_en | 1,0004995 | 0,9998288 | 1,0011173 | 0,1255318 | 0,0004994 | 0,000326 |
| Anx past month | Fibre_en | 0,9794407 | 0,9557065 | 1,0032399 | 0,09333 | -0,0207735 | 0,0123793 |
| Anx past month | Calcium_en | 0,9505294 | 0,6361948 | 1,4031498 | 0,8013521 | -0,0507362 | 0,2016562 |
| **Anx past month** | **Omega3_en** | **0,9988776** | **0,998055** | **0,9996308** | **0,0052358** | **-0,001123** | **0,0004022** |
| Anx past month | PUFA_en | 0,9977651 | 0,9345154 | 1,0624898 | 0,9454663 | -0,0022374 | 0,03271 |
| Anx past month | Trans_fat_en | 1,1074715 | 0,3533265 | 3,4124007 | 0,8598642 | 0,1020795 | 0,5782002 |
| **Model 2: adjusted for sex, age, education** | | | |  |  |  |  |
| **Outcome** | **Food** | **OR** | **95%CI_lo** | **95%CI_hi** | **p** | **B** | **SE** |
| Anx past month | Fruit_en | 0,9993905 | 0,9983132 | 1,0004237 | 0,2573458 | -0,0006097 | 0,0005382 |
| **Anx past month** | **Vegetables_en** | **0,9977337** | **0,9961469** | **0,9992573** | **0,004306** | **-0,0022689** | **0,0007947** |
| Anx past month | Legumes_en | 1,0008122 | 0,9957494 | 1,0053793 | 0,7395623 | 0,0008119 | 0,0024422 |
| Anx past month | Wholegrains_en | 0,9994084 | 0,9972217 | 1,0015729 | 0,594008 | -0,0005917 | 0,0011101 |
| Anx past month | Nuts_seeds_en | 1,0005644 | 0,9927371 | 1,0078393 | 0,8832835 | 0,0005643 | 0,0038436 |
| Anx past month | Milk_en | 1,0002425 | 0,9995533 | 1,0008864 | 0,4737408 | 0,0002425 | 0,0003384 |
| **Anx past month** | **Red_meat_en** | **0,9941974** | **0,9893131** | **0,9989287** | **0,01834** | **-0,0058195** | **0,0024673** |
| Anx past month | Processed_meat_en | 1,000915 | 0,9920318 | 1,0093698 | 0,8357357 | 0,0009146 | 0,0044109 |
| Anx past month | Sweet_drinks_en | 1,0003239 | 0,9996242 | 1,0009711 | 0,3421664 | 0,0003238 | 0,0003409 |
| Anx past month | Fibre_en | 0,9849319 | 0,9604324 | 1,0095066 | 0,2322054 | -0,0151828 | 0,0127085 |
| Anx past month | Calcium_en | 0,9592135 | 0,6364957 | 1,4263576 | 0,839559 | -0,0416416 | 0,2056804 |
| **Anx past month** | **Omega3_en** | **0,9990384** | **0,9982055** | **0,9997987** | **0,0179603** | **-0,0009621** | **0,0004066** |
| Anx past month | PUFA_en | 1,0014235 | 0,9372933 | 1,0670873 | 0,9656698 | 0,0014224 | 0,0330493 |
| Anx past month | Trans_fat_en | 0,8787343 | 0,2755466 | 2,7541925 | 0,8256582 | -0,1292727 | 0,5868735 |
| **Model 3: adjusted for sex, age, education, smoking, body mass index and physical activity*** | | | | | | | |
| **Outcome** | **Food** | **OR** | **95%CI_lo** | **95%CI_hi** | **p** | **B** | **SE** |
| Anx past month | Fruit_en | 0,999897 | 0,9987031 | 1,0010398 | 0,8626226 | -0,000103 | 0,0005952 |
| **Anx past month** | **Vegetables_en** | **0,9979304** | **0,996118** | **0,9996643** | **0,0222031** | **-0,0020718** | **0,0009059** |
| Anx past month | Legumes_en | 1,0003745 | 0,9943078 | 1,0058043 | 0,8978466 | 0,0003744 | 0,0029161 |
| Anx past month | Wholegrains_en | 0,9998788 | 0,9974394 | 1,0022903 | 0,9218925 | -0,0001213 | 0,0012367 |
| Anx past month | Nuts_seeds_en | 1,0002558 | 0,9913433 | 1,0084243 | 0,9530898 | 0,0002557 | 0,0043471 |
| Anx past month | Milk_en | 1,0002037 | 0,9994171 | 1,0009434 | 0,5998373 | 0,0002037 | 0,0003883 |
| **Anx past month** | **Red_meat_en** | **0,9915503** | **0,9858491** | **0,9970442** | **0,0032252** | **-0,0084856** | **0,0028809** |
| Anx past month | Processed_meat_en | 1,0015557 | 0,9918503 | 1,0107551 | 0,7460961 | 0,0015545 | 0,0048009 |
| Anx past month | Sweet_drinks_en | 1,0001134 | 0,9992589 | 1,0008817 | 0,7824845 | 0,0001134 | 0,0004107 |
| Anx past month | Fibre_en | 0,9871398 | 0,959028 | 1,0153733 | 0,373864 | -0,0129436 | 0,0145555 |
| Anx past month | Calcium_en | 1,0233693 | 0,6394841 | 1,6161533 | 0,9221171 | 0,0231004 | 0,2362796 |
| Anx past month | Omega3_en | 0,9992027 | 0,998298 | 1,0000175 | 0,0689646 | -0,0007976 | 0,0004386 |
| Anx past month | PUFA_en | 1,0236754 | 0,9504716 | 1,0992026 | 0,5274086 | 0,0233995 | 0,0370264 |
| Anx past month | Trans_fat_en | 0,8137348 | 0,2124411 | 3,0484046 | 0,7614124 | -0,2061208 | 0,6788626 |
| Abbreviations: Anx: Anxiety Disorder, OR: Odds ratio, CI: confidence interval, SE: standard error | | | | | | | |
| *n=1338 |  |  |  |  |  |  |  |

| **Supplemental table 3. Logistic regression on the association of food groups with QIDS depression, Willet adjusted model (n=1615)** | | | | | | | |
| --- | --- | --- | --- | --- | --- | --- | --- |
| **Model 1: Univariable** | |  |  |  |  |  |  |
| **Outcome** | **Food** | **OR** | **95%CI_lo** | **95%CI_hi** | **p** | **B** | **SE** |
| QIDS>=13 | Fruit_en | 0,9986381 | 0,9971706 | 1,0000174 | 0,0608006 | -0,0013628 | 0,0007269 |
| **QIDS>=13** | **Vegetables_en** | **0,9969895** | **0,9949216** | **0,9989435** | **0,0033785** | **-0,0030151** | **0,0010287** |
| QIDS>=13 | Legumes_en | 0,9992098 | 0,992326 | 1,0050561 | 0,8072367 | -0,0007905 | 0,0032399 |
| **QIDS>=13** | **Wholegrains_en** | **0,9952995** | **0,9924207** | **0,9981293** | **0,0012827** | **-0,0047116** | **0,0014633** |
| QIDS>=13 | Nuts_seeds_en | 0,9970555 | 0,9865955 | 1,0064275 | 0,5613263 | -0,0029488 | 0,0050765 |
| QIDS>=13 | Milk_en | 1,0002533 | 0,9993613 | 1,0010487 | 0,5548624 | 0,0002532 | 0,0004289 |
| QIDS>=13 | Red_meat_en | 1,0022382 | 0,9966041 | 1,0075862 | 0,4235194 | 0,0022357 | 0,0027935 |
| QIDS>=13 | Processed_meat_en | 0,9973575 | 0,9856164 | 1,0081218 | 0,645626 | -0,002646 | 0,0057542 |
| QIDS>=13 | Sweet_drinks_en | 1,0006224 | 0,9997911 | 1,0013495 | 0,1131693 | 0,0006222 | 0,0003928 |
| **QIDS>=13** | **Fibre_en** | **0,9420542** | **0,9111541** | **0,9731786** | **0,0003809** | **-0,0596925** | **0,0168008** |
| QIDS>=13 | Calcium_en | 0,7768924 | 0,4536492 | 1,2991668 | 0,3469835 | -0,2524534 | 0,2684371 |
| QIDS>=13 | Omega3_en | 0,9989888 | 0,9979121 | 0,9999418 | 0,0508737 | -0,0010117 | 0,0005181 |
| QIDS>=13 | PUFA_en | 0,98197 | 0,9003096 | 1,0653945 | 0,6717275 | -0,0181945 | 0,042934 |
| **QIDS>=13** | **Trans_fat_en** | **8,0463685** | **1,9521066** | **32,428479** | **0,0035853** | **2,0852209** | **0,715951** |
| **Model 2: adjusted for sex, age, education** | | | |  |  |  |  |
| **Outcome** | **Food** | **OR** | **95%CI_lo** | **95%CI_hi** | **p** | **B** | **SE** |
| QIDS>=13 | Fruit_en | 0,9986934 | 0,9972277 | 1,0000672 | 0,0712616 | -0,0013075 | 0,0007248 |
| **QIDS>=13** | **Vegetables_en** | **0,9974441** | **0,9953299** | **0,9994435** | **0,0149449** | **-0,0025592** | **0,0010515** |
| QIDS>=13 | Legumes_en | 1,0001582 | 0,9932516 | 1,0060087 | 0,9610393 | 0,0001582 | 0,0032382 |
| **QIDS>=13** | **Wholegrains_en** | **0,9957967** | **0,992895** | **0,9986545** | **0,0042965** | **-0,0042122** | **0,0014751** |
| QIDS>=13 | Nuts_seeds_en | 0,9993456 | 0,9889222 | 1,0086303 | 0,8963769 | -0,0006546 | 0,0050263 |
| QIDS>=13 | Milk_en | 1,0001713 | 0,9992615 | 1,0009821 | 0,6954575 | 0,0001713 | 0,0004377 |
| QIDS>=13 | Red_meat_en | 1,0013204 | 0,9955077 | 1,0068598 | 0,648087 | 0,0013196 | 0,0028911 |
| QIDS>=13 | Processed_meat_en | 0,9936432 | 0,9813927 | 1,0049564 | 0,2919393 | -0,0063771 | 0,0060511 |
| QIDS>=13 | Sweet_drinks_en | 1,0006344 | 0,9997804 | 1,0013903 | 0,1182095 | 0,0006342 | 0,000406 |
| **QIDS>=13** | **Fibre_en** | **0,9483389** | **0,9165766** | **0,9803212** | **0,0019849** | **-0,0530433** | **0,0171524** |
| QIDS>=13 | Calcium_en | 0,7866869 | 0,455172 | 1,3242024 | 0,3784122 | -0,2399249 | 0,2723864 |
| QIDS>=13 | Omega3_en | 0,9990679 | 0,9979698 | 1,0000358 | 0,0770224 | -0,0009325 | 0,0005274 |
| QIDS>=13 | PUFA_en | 0,9900022 | 0,9080311 | 1,0737183 | 0,8140896 | -0,0100481 | 0,0427299 |
| **QIDS>=13** | **Trans_fat_en** | **6,1092672** | **1,4718172** | **24,875843** | **0,0119679** | **1,8098068** | **0,7201522** |
| **Model 3: adjusted for sex, age, education, smoking, body mass index and physical activity*** | | | | | | | |
| **Outcome** | **Food** | **OR** | **95%CI_lo** | **95%CI_hi** | **p** | **B** | **SE** |
| QIDS>=13 | Fruit_en | 0,9988025 | 0,9971395 | 1,0003449 | 0,142923 | -0,0011982 | 0,0008179 |
| **QIDS>=13** | **Vegetables_en** | **0,9976491** | **0,9952641** | **0,9999019** | **0,0470275** | **-0,0023537** | **0,0011851** |
| QIDS>=13 | Legumes_en | 1,0002743 | 0,992399 | 1,0070038 | 0,9408139 | 0,0002743 | 0,0036945 |
| QIDS>=13 | Wholegrains_en | 0,9973218 | 0,99415 | 1,000455 | 0,0959476 | -0,0026818 | 0,0016109 |
| QIDS>=13 | Nuts_seeds_en | 0,9968848 | 0,9847111 | 1,0076987 | 0,5957243 | -0,00312 | 0,0058806 |
| QIDS>=13 | Milk_en | 1,000007 | 0,9989506 | 1,0009712 | 0,9891058 | 7,019E-06 | 0,000514 |
| QIDS>=13 | Red_meat_en | 1,0000198 | 0,9935893 | 1,0060897 | 0,9950503 | 1,977E-05 | 0,0031861 |
| QIDS>=13 | Processed_meat_en | 0,9955372 | 0,9826482 | 1,0073111 | 0,47867 | -0,0044728 | 0,0063135 |
| QIDS>=13 | Sweet_drinks_en | 0,9999425 | 0,9987655 | 1,000926 | 0,9160159 | -5,752E-05 | 0,0005455 |
| **QIDS>=13** | **Fibre_en** | **0,9558089** | **0,9192847** | **0,9923501** | **0,0204823** | **-0,0451973** | **0,0195035** |
| QIDS>=13 | Calcium_en | 0,7255803 | 0,3864689 | 1,3263017 | 0,3074143 | -0,3207836 | 0,3142895 |
| QIDS>=13 | Omega3_en | 0,9994086 | 0,99828 | 1,0003863 | 0,2701734 | -0,0005916 | 0,0005365 |
| QIDS>=13 | PUFA_en | 0,9912773 | 0,8997977 | 1,0854212 | 0,8544908 | -0,0087609 | 0,0477718 |
| **QIDS>=13** | **Trans_fat_en** | **8,3470953** | **1,6471041** | **42,214161** | **0,0101543** | **2,1219136** | **0,8254771** |
| Abbreviations: QIDS: Quick Inventory of Depressive symptomatology >=13, OR: Odds ratio, CI: confidence interval, SE: standard error | | | | | | | |
| *smaller n due to missing values for lifestyle variables | | | | |  |  |  |

| **Supplemental Table 4. Logistic regression on the association of food groups with BAI anxiety, Willet adjusted model (n=1612)** | | | | | | | |
| --- | --- | --- | --- | --- | --- | --- | --- |
| **Model 1: Univariable** | |  |  |  |  |  |  |
| **Outcome** | **Food** | **OR** | **95%CI_lo** | **95%CI_hi** | **p** | **B** | **SE** |
| BAI>=16 | Fruit_en | 0,9997227 | 0,9986754 | 1,000729 | 0,5962643 | -0,0002774 | 0,0005236 |
| **BAI>=16** | **Vegetables_en** | **0,9974863** | **0,9959378** | **0,9989718** | **0,0011727** | **-0,0025168** | **0,0007755** |
| BAI>=16 | Legumes_en | 0,9986195 | 0,9932655 | 1,0034093 | 0,5929455 | -0,0013814 | 0,0025842 |
| **BAI>=16** | **Wholegrains_en** | **0,9972358** | **0,9950427** | **0,9993927** | **0,0128473** | **-0,002768** | **0,0011126** |
| BAI>=16 | Nuts_seeds_en | 0,9949938 | 0,9867867 | 1,0026001 | 0,2156992 | -0,0050188 | 0,0040538 |
| BAI>=16 | Milk_en | 0,9999724 | 0,9992522 | 1,0006388 | 0,9378033 | -2,755E-05 | 0,0003531 |
| BAI>=16 | Red_meat_en | 1,0012983 | 0,9968816 | 1,0055774 | 0,5576958 | 0,0012975 | 0,0022131 |
| BAI>=16 | Processed_meat_en | 1,003395 | 0,9950046 | 1,0113803 | 0,4145653 | 0,0033893 | 0,0041541 |
| BAI>=16 | Sweet_drinks_en | 1,0003185 | 0,9996168 | 1,0009551 | 0,3472171 | 0,0003185 | 0,0003388 |
| **BAI>=16** | **Fibre_en** | **0,9711392** | **0,947222** | **0,9951216** | **0,0199327** | **-0,0292854** | **0,0125817** |
| BAI>=16 | Calcium_en | 0,6831213 | 0,4485843 | 1,0261362 | 0,0709405 | -0,3810828 | 0,2110259 |
| BAI>=16 | Omega3_en | 0,9993266 | 0,9985648 | 1,0000244 | 0,0702692 | -0,0006737 | 0,0003722 |
| BAI>=16 | PUFA_en | 0,9959339 | 0,9326059 | 1,0607184 | 0,9011586 | -0,0040744 | 0,0328058 |
| BAI>=16 | Trans_fat_en | 1,7366643 | 0,5552343 | 5,3494366 | 0,3391805 | 0,5519662 | 0,5775 |
| **Model 2: adjusted for sex, age, education** | | | |  |  |  |  |
| **Outcome** | **Food** | **OR** | **95%CI_lo** | **95%CI_hi** | **p** | **B** | **SE** |
| BAI>=16 | Fruit_en | 0,9997816 | 0,9987333 | 1,0007898 | 0,6767996 | -0,0002185 | 0,0005241 |
| **BAI>=16** | **Vegetables_en** | **0,9979172** | **0,9963296** | **0,9994421** | **0,0087362** | **-0,002085** | **0,0007951** |
| BAI>=16 | Legumes_en | 0,9998967 | 0,9945196 | 1,0047121 | 0,9681709 | -0,0001033 | 0,0025897 |
| BAI>=16 | Wholegrains_en | 0,9978245 | 0,9955871 | 1,0000328 | 0,0552388 | -0,0021779 | 0,0011361 |
| BAI>=16 | Nuts_seeds_en | 0,9971854 | 0,9888948 | 1,0048553 | 0,4896915 | -0,0028185 | 0,0040801 |
| BAI>=16 | Milk_en | 0,9998811 | 0,9991448 | 1,0005613 | 0,7417819 | -0,0001189 | 0,0003608 |
| BAI>=16 | Red_meat_en | 1,0005387 | 0,9959491 | 1,0049919 | 0,8151623 | 0,0005386 | 0,0023039 |
| BAI>=16 | Processed_meat_en | 1,0010627 | 0,9922603 | 1,0094538 | 0,808075 | 0,0010622 | 0,0043727 |
| BAI>=16 | Sweet_drinks_en | 1,0002154 | 0,9994891 | 1,0008788 | 0,5404399 | 0,0002154 | 0,0003519 |
| BAI>=16 | Fibre_en | 0,9788659 | 0,9540892 | 1,0037336 | 0,0986942 | -0,0213607 | 0,0129363 |
| BAI>=16 | Calcium_en | 0,6747876 | 0,4379984 | 1,0237033 | 0,0692118 | -0,3933573 | 0,2164831 |
| BAI>=16 | Omega3_en | 0,9994671 | 0,9986938 | 1,0001744 | 0,1576597 | -0,0005331 | 0,0003773 |
| BAI>=16 | PUFA_en | 1,0028478 | 0,9387299 | 1,0685058 | 0,931321 | 0,0028437 | 0,0329964 |
| BAI>=16 | Trans_fat_en | 1,2525733 | 0,3937551 | 3,9229373 | 0,7007231 | 0,2252 | 0,5859327 |
| **Model 3: adjusted for sex, age, education, smoking, body mass index and physical activity*** | | | | | | | |
| **Outcome** | **Food** | **OR** | **95%CI_lo** | **95%CI_hi** | **p** | **B** | **SE** |
| BAI>=16 | Fruit_en | 1,0002075 | 0,9990346 | 1,0013358 | 0,7230796 | 0,0002075 | 0,0005856 |
| BAI>=16 | Vegetables_en | 0,9986196 | 0,9968297 | 1,0003389 | 0,1229902 | -0,0013813 | 0,0008956 |
| BAI>=16 | Legumes_en | 0,9995316 | 0,9931599 | 1,0052334 | 0,878545 | -0,0004685 | 0,0030658 |
| BAI>=16 | Wholegrains_en | 1,000151 | 0,9976183 | 1,002672 | 0,9066625 | 0,000151 | 0,0012879 |
| BAI>=16 | Nuts_seeds_en | 0,995437 | 0,9857368 | 1,0043514 | 0,3375012 | -0,0045735 | 0,0047684 |
| BAI>=16 | Milk_en | 0,9997141 | 0,9988432 | 1,0005267 | 0,5047871 | -0,000286 | 0,0004288 |
| BAI>=16 | Red_meat_en | 0,9997232 | 0,9944993 | 1,0047521 | 0,9156261 | -0,0002768 | 0,002613 |
| BAI>=16 | Processed_meat_en | 1,0027117 | 0,9931363 | 1,0118501 | 0,5680104 | 0,002708 | 0,0047427 |
| BAI>=16 | Sweet_drinks_en | 0,9998656 | 0,9989327 | 1,000687 | 0,7621236 | -0,0001344 | 0,000444 |
| BAI>=16 | Fibre_en | 0,9941496 | 0,9653194 | 1,0231176 | 0,6921862 | -0,0058676 | 0,0148214 |
| **BAI>=16** | **Calcium_en** | **0,584416** | **0,3496057** | **0,959414** | **0,0368595** | **-0,5371422** | **0,2573362** |
| BAI>=16 | Omega3_en | 0,9997703 | 0,9989612 | 1,0005049 | 0,5584989 | -0,0002297 | 0,0003926 |
| BAI>=16 | PUFA_en | 1,0114519 | 0,9377453 | 1,0875968 | 0,7629511 | 0,0113869 | 0,0377539 |
| BAI>=16 | Trans_fat_en | 1,2449899 | 0,3261306 | 4,6756956 | 0,746705 | 0,2191274 | 0,6784401 |
| Abbreviations: BAI: Becks Anxiety Index >=16, OR: Odds ratio, CI: confidence interval, SE: standard error | | | | | | | |
| *smaller n due to missing values for lifestyle variables | | | | |  |  |  |

**Supplemental table 5. Abbreviations of food groups in regression models**

| **Variable name** | **Food group** |
| --- | --- |
| Fruit | Fruit g/day |
| Vegetables | Vegetables g/day |
| Legumes | Legumes g/day |
| Wholegrains | Whole grains g/day |
| Nuts_seeds | Nuts and seeds g/day |
| Milk | Milk g/day |
| Red_meat | Red meat g/day |
| Processed_meat | Processed meat g/day |
| Sweet_drinks | Sweet drinks g/day |
| Fibre | Fibre g/day |
| Calcium | Calcium g/day |
| Omega3 | Omega-3 (DHA and EPA) mg/day |
| PUFA | PUFA (% energy) |
| Trans_fat | Transfat (% energy) |

| **Variable name** | **Food group** |
| --- | --- |
| Fruit_en | Fruit g/day, energy-adjusted Willett method |
| Vegetables_en | Vegetables g/day, energy-adjusted Willett method |
| Legumes_en | Legumes g/day, energy-adjusted Willett method |
| Wholegrains_en | Whole grains g/day, energy-adjusted Willett method |
| Nuts_seeds_en | Nuts and seeds g/day, energy-adjusted Willett method |
| Milk_en | Milk g/day, energy-adjusted Willett method |
| Red_meat_en | Red meat g/day, energy-adjusted Willett method |
| Processed_meat_en | Processed meat g/day, energy-adjusted Willett method |
| Sweet_drinks_en | Sweet drinks g/day, energy-adjusted Willett method |
| Fibre_en | Fibre g/day, energy-adjusted Willett method |
| Calcium_en | Calcium g/day, energy-adjusted Willett method |
| Omega3_en | Omega-3 (DHA and EPA) mg/day, energy-adjusted Willett method |
| PUFA_en | PUFA (% energy), energy-adjusted Willett method |
| Trans_fat_en | Transfat (% energy), energy-adjusted Willett method |

**Supplemental table 6. Sensitivity analysis with natural logarithm transformed PUFA**

| **Supplemental Table 6A. Logistic regression on the association of LN-transformed PUFA with MDD in the past month (n=1634)** | | | | | | | | | | | | |
| --- | --- | --- | --- | --- | --- | --- | --- | --- | --- | --- | --- | --- |
| **Model 1: Univariable** | | |  | |  | |  | |  | |  | |
| **Outcome** | **Food** | **OR** | | **95%CI_lo** | | **95%CI_hi** | | **p** | | **B** | | **SE** |
| MDD past month | LN_PUFA | 0,553432 | | 0,311903 | | 0,981995 | | 0,04317 | | -0,59162 | | 0,292581 |
| **Model 2: adjusted for sex, age, education** | | | | | | |  | |  | |  | |
| **Outcome** | **Food** | **OR** | | **95%CI_lo** | | **95%CI_hi** | | **p** | | **B** | | **SE** |
| MDD past month | LN_PUFA | 0,57916 | | 0,325459 | | 1,030624 | | 0,063257 | | -0,54618 | | 0,294057 |
| **Model 3: adjusted for sex, age, education, smoking, body mass index and physical activity*** | | | | | | | | | | | | |
| **Outcome** | **Food** | **OR** | | **95%CI_lo** | | **95%CI_hi** | | **p** | | **B** | | **SE** |
| MDD past month | LN_PUFA | 0,489199 | | 0,255674 | | 0,936022 | | 0,030798 | | -0,71499 | | 0,331062 |
| Abbreviations: MDD: Major Depressive Disoder, OR: Odds ratio, CI: confidence interval, SE: standard error | | | | | | | | | | | | |
| *n=1338 |  |  | |  | |  | |  | |  | |  |
| **Supplemental Table 6B. Logistic regression on the association of LN-transformed PUFA with anxiety in the past month (n=1634)** | | | | | | | | | | | | |
| **Model 1: Univariable** | | |  | |  | |  | |  | |  | |
| **Outcome** | **Food** | **OR** | | **95%CI_lo** | | **95%CI_hi** | | **p** | | **B** | | **SE** |
| Anx past month | LN_PUFA | 0,938739 | | 0,593414 | | 1,485017 | | 0,787041 | | -0,06322 | | 0,234007 |
| **Model 2: adjusted for sex, age, education** | | | | | | |  | |  | |  | |
| **Outcome** | **Food** | **OR** | | **95%CI_lo** | | **95%CI_hi** | | **p** | | **B** | | **SE** |
| Anx past month | LN_PUFA | 0,971112 | | 0,610432 | | 1,544903 | | 0,901513 | | -0,02931 | | 0,236879 |
| **Model 3: adjusted for sex, age, education, smoking, body mass index and physical activity*** | | | | | | | | | | | | |
| **Outcome** | **Food** | **OR** | | **95%CI_lo** | | **95%CI_hi** | | **p** | | **B** | | **SE** |
| Anx past month | LN_PUFA | 1,083449 | | 0,640385 | | 1,833059 | | 0,765135 | | 0,08015 | | 0,268289 |
| Abbreviations: Anx: Anxiety disorder, OR: Odds ratio, CI: confidence interval, SE: standard error | | | | | | | | | | | | |
| *n=1338 |  |  | |  | |  | |  | |  | |  |
| **Supplemental Table 6C. Logistic regression on the association of LN-transformed PUFA with QIDS depression (n=1615)** | | | | | | | | | | | | |
| **Model 1: Univariable** | | |  | |  | |  | |  | |  | |
| **Outcome** | **Food** | **OR** | | **95%CI_lo** | | **95%CI_hi** | | **p** | | **B** | | **SE** |
| QIDS | LN_PUFA | 0,686963 | | 0,379938 | | 1,242094 | | 0,214041 | | -0,37547 | | 0,302186 |
| **Model 2: adjusted for sex, age, education** | | | | | | |  | |  | |  | |
| **Outcome** | **Food** | **OR** | | **95%CI_lo** | | **95%CI_hi** | | **p** | | **B** | | **SE** |
| QIDS | LN_PUFA | 0,741054 | | 0,409984 | | 1,339469 | | 0,321077 | | -0,29968 | | 0,302024 |
| **Model 3: adjusted for sex, age, education, smoking, body mass index and physical activity*** | | | | | | | | | | | | |
| **Outcome** | **Food** | **OR** | | **95%CI_lo** | | **95%CI_hi** | | **p** | | **B** | | **SE** |
| QIDS | LN_PUFA | 0,780677 | | 0,404117 | | 1,508119 | | 0,461129 | | -0,24759 | | 0,335954 |
| Abbreviations: QIDS: Quick Inventory of Depressive Symptomatology, OR: Odds ratio, CI: confidence interval, SE: standard error | | | | | | | | | | | | |
| *smaller n due to missings in lifestyle variables | | | | | | |  | |  | |  | |
| **Supplemental Table 6D. Logistic regression on the association of LN-transformed PUFA with BAI anx (n=1612)** | | | | | | | | | | | | |
| **Model 1: Univariable** | | |  | |  | |  | |  | |  | |
| **Outcome** | **Food** | **OR** | | **95%CI_lo** | | **95%CI_hi** | | **p** | | **B** | | **SE** |
| BAI | LN_PUFA | 0,886089 | | 0,559401 | | 1,403562 | | 0,606313 | | -0,12094 | | 0,234673 |
| **Model 2: adjusted for sex, age, education** | | | | | | |  | |  | |  | |
| **Outcome** | **Food** | **OR** | | **95%CI_lo** | | **95%CI_hi** | | **p** | | **B** | | **SE** |
| BAI | LN_PUFA | 0,946325 | | 0,595025 | | 1,505032 | | 0,815726 | | -0,05517 | | 0,236731 |
| **Model 3: adjusted for sex, age, education, smoking, body mass index and physical activity*** | | | | | | | | | | | | |
| **Outcome** | **Food** | **OR** | | **95%CI_lo** | | **95%CI_hi** | | **p** | | **B** | | **SE** |
| BAI | LN_PUFA | 1,004682 | | 0,592861 | | 1,702566 | | 0,986153 | | 0,004671 | | 0,26912 |
| Abbreviations: BAI: Becks Anxiety Index, OR: Odds ratio, CI: confidence interval, SE: standard error | | | | | | | | | | | | |
| *smaller n due to missings in lifestyle variables | | | | | | |  | |  | |  | |
